# Early Epidemiological Evidence of Public Health Value of WA Notify, a Smartphone-based Exposure Notification Tool: Modeling COVID-19 Cases Averted in Washington State

**DOI:** 10.1101/2021.06.04.21257951

**Authors:** Courtney Segal, Zhehao Zhang, Bryant T Karras, Debra Revere, Gregory Zane, Janet G Baseman

## Abstract

**Background:** Secure and anonymous smartphone-based exposure notification tools are recently developed public health interventions that aim to reduce COVID-19 transmission and supplement traditional public health suriveillance. We assessed the impact of Washington State’s exposure notification tool, WA Notify, in mitigating the spread of COVID-19 during its first four months of implementation.

**Methods:** Due to the constraints of privacy-preservation, aggregate metrics and disparate data sources were utilized to estimate the number of COVID-19 cases averted based on a modelling approach adapted from Wymant et al (2021) using the following parameters: number of notifications generated; the probability that a notified individual goes on to become a case; expected fraction of transmissions preventable by strict quarantine after notification; actual adherence to quarantine; and expected size of the full transmission chain if a contact had not been notified.

**Results:** The model was run on a range of secondary attack rates (5.1%-13.706%) and quarantine effectiveness (53% and 64%). Assuming a 12.085% secondary attack rate and 53% quarantine effectiveness, the model shows that 5,500 cases (central 95% range of sensitivity analyses 2,800-8,200) were averted statewide during the first four months of its implementation. Based on an estimated COVID-19 case fatality of 1.4%, WA Notify saved 40-115 lives during the study period.

**Conclusions:** These findings demonstrate the value of exposure notification tools as a novel public health intervention to mitigate the spread of COVID-19 in the U.S. As new variants emerge and non-essential travel bans are lifted, exposure notification tools may continue to play a valuable role in limiting the spread of COVID-19.

## Background

Case investigation and contact tracing (CI/CT) are core, evidence-based strategies employed by public health agencies (PHAs) to control the spread of infectious diseases^1^. By conducting interviews with positive index cases, potentially exposed contacts are identified, subsequently informed of their exposure and provided guidance regarding testing, symptom monitoring and recommendations for engagment in protective measures^2^. This process has successfully limited forward disease transmission in prior epidemics^3,4^ despite its challenges: scalability, contact notification delay, challenges to recall contacts and the existence of unknown, unidentifiable contacts^5,6^. With the rapid spread of severe acute respiratory syndrome coronavirus-2 (SARS-CoV-2) overwhelming CI/CT capacity in many parts of the U.S.^7^, smartphone-based exposure notification (EN) tools have been developed and utilized in some U.S. states as a supplemental strategy to traditional PHA CI/CT processes^8^.

In May 2020, the Google|Apple protocol GAEN (Google-Apple Exposure Notifications) was released for use by PHAs within their jurisdictions. A GAEN-based tool is considered “privacy-preserving” in that user’s phones log their exposure history without requiring geospatial location tracking or allowing access to personal data. GAEN-based tools operate as decentralized systems. Rather than have each State and Territory build and host its own verification and key servers, in the U.S., the Association of Public Health Laboratories (APHL) hosts both a multi-tenant verification (Google) and a key server (Microsoft) to support GAEN implementations. Once added to a smartphone, EN tools utilize Bluetooth technology to determine digital proximity between devices and exchange random, ephemeral cryptographic keys when two smartphones are within the physical distance and duration of time specified by a given PHA^9^. A user who tests positive for COVID-19 can anonymously report their diagnosis through the tool by voluntarily entering a verification code which then uploads keys to the APHL server^10^. Other users whose proximity and duration of exposure to the index case match specifications set by the PHA are alerted with an EN message. WA Notify^11^, using the GAEN EN Express solution launched in Washington (WA) State, has been installed on more than two million devices, representing approximately 33% of the adult population. A detailed overview of the WA Notify user engagement experience and workflow (Figure S-1) can be found in the *Supplementary Appendix*.

EN technology represents a potentially disruptive public health strategy for pandemic control. Recent modelling and simulation evaluations have demonstrated the epidemiological value of EN tools in improving contact identification, particularly of contacts unknown to the index case, and slowing secondary transmission of infection^12–15^. In January 2021, the University of Washington (UW), in partnership with the WA State Department of Health (DOH), conducted an evaluation of WA Notify’s effectiveness in mitigating the spread of COVID-19 during its first four months of implementation: 11/30/2020—03/31/2021. This timeframe (see Figures S-2 and S-3 in *Appendix*) represents the peak of COVID-19 case surges and the highest volume of CI/CT activity since the start of the pandemic, with 196,036 new cases, 8,571 hospitalizations, and 2,464 COVID-related deaths documented by WA DOH^16^.

The preliminary evaluation of WA Notify described here seeks to answer the question: To what extent did WA Notify avert new COVID-19 cases in Washington during the first four months of its use?

## Methods

Empirical validation of WA Notify’s epidemiological impact and value as a non-pharmaceutical intervention (NPI) is limited by the anonymity of users and data privacy. Within these constraints, the approach described herein were designed to leverage aggregate metrics across disparate sources^17,18^. The UW Institutional Review Board reviewed the evaluation plan, and determined it was a public health quality improvement project and non-human subjects research.

### Data Sources

A brief summary of the data sources used for this analysis is provided below. Please refer to Table S-1 in the *Supplementary Appendix* for a detailed description of all data sources used in this evaluation.

- DOH CI/CT Data. De-identified dataset (n=18,616 index case investigation records linked to at least one close contact; n=49,488 completed contact tracing interviews) representing an average of 2.7 contacts per index case.
- APHL EN Verification Code Server Metrics. Number of codes deployed, claimed, and the distribution of the time from code issued to code claimed. WA State initially issued codes manually by phone and switched to a bulk texting protocol on 01/11/2021; a total of 101,990 codes were issued with 10,084 claimed (9.9%), representing 5.1% of all positive cases during the study period (see Figure S-4 in *Appendix*). These data, along with specimen collection date from the CI/CT dataset, were used to estimate the time delay from lab reporting to code issue.
- Landing Page pagehits. Accessible by tapping the “What next?” link in a WA Notify EN message, DOH’s Landing Page provides information about what to do after learning of a potential close contact exposure to an index case. Between 11/30/2020—03/31/2021, there were 16,748 Landing Page hits, representing the subset of EN recipients who tapped the link. The Landing Page also hosts anonymous surveys regarding protective behaviors.
- Protective Behavior Surveys. Responses to Survey 1 (N=1,132) were used to estimate quarantine behavior after receiving an EN (e.g. avoiding public places, and/or staying away from others in their household). Survey 2 (N=219), a follow-up 2 weeks later, captured self-report of actual quarantine behavior after EN receipt. Forty-two percent of Survey 1 respondents (N=475) reported intent to quarantine and 64% of Survey 2 respondents (N=140) reported having engaged in some form of quarantine behavior after receiving an EN.
- Exposure Notifications Private Analytics (ENPA) Dashboard Data. Available beginning 02/09/2021, the dashboard provides aggregated, anonymous data regarding ENs received, opened, or dismissed. An estimated 10,741 ENs were generated and 5,215 ENs opened among the sample of WA Notify users who opted-in to share their analytics (representing approximately 20% of all users).

### Modeling Approach and Parameter Estimation

To estimate the number of cases averted by WA Notify, a modeling approach developed to evaluate the NHS COVID-19 app was adapted^12^. This model is essentially a product of five terms: (i) number of notifications generated, (ii) secondary attack rate, i.e., the probability that notified individuals go on to become cases, (iii) expected fraction of transmissions preventable by strict quarantine of an infectious individual after a notification, (iv) actual adherence to quarantine, and (v) expected size of the full transmission chain that would be originated by the contact if they had not been notified.

#### Number of notifications generated (i)

The precise number of notifications generated and received is not available due to differential privacy methods^19^. Daily Landing Page page hits and aggregate ENPA Dashboard Data for the month of March were used to estimate the total number of notifications generated. Using the proportion of ENs opened of those generated, we assume a 1/2.06 or 48.54% open ratio for notifications. With a total of 16,748 landing page hits between 11/30 to 3/31, we estimate that 16748*2.06 = 34,501 ENs were generated. A Pearsons correlation test determined the estimate was a good fit (see Fig. S-5 in the *Appendix*).

#### Secondary attack rate (ii)

Secondary attack rate (SAR) refers to the extent to which person-to-person spread of an infectious illness occurs from an index case^20^. We estimated SAR using the CI/CT and ENPA data to reflect different real-world scenarios of WA Notify users, as follows:

Using CI/CT data, 12,794 of 56,926 reported close contacts were non-household contacts, i.e., a contact meeting exposure criteria who does not live in the same residence as the index case. Of these contacts, 852 were diagnosed with COVID-19 during the study period, resulting in an *out-of-household SAR* of 6.659%. For household contacts, i.e., a contact meeting exposure criteria who lives in or shares the same residence as an index case, 5868 of 42,813 household contacts were confirmed cases, resulting in an *in-household SAR* of 13.706%. The *overall upperbound SAR estimate* derived from the CI/CT dataset is the weighted average of the *out-of-household* and *in-household* SAR estimates, (852+5868)/(42813+12794) = 12.085%.

Using the ENPA Dashboard data, we estimated the spread of COVID-19 within the WA Notify population as the proportion of users who claimed a code and *also* received an EN (N=552) among the notifications generated (N=10,741), which provided a *lower bound SAR estimate* of 5.1%. The propagation of standard deviations for this estimate is reported in Section C of the *Appendix*.

#### Expected fraction of transmissions prevented (iii)

The expected fraction of transmissions preventable by strict quarantine of an infectious individual after a notification depends on the delay from the time of exposure to a positive COVID-19 index case to the time of EN receipt^21^. Estimating this time delay requires integrating estimates from multiple real-world data sources, as illustrated in Fig. 1. A detailed description of methods and outcomes of these time calculations can be found in the *Appendix* (see Figs. S-6, S-7, S-8, S-9).

**Figure 1.**
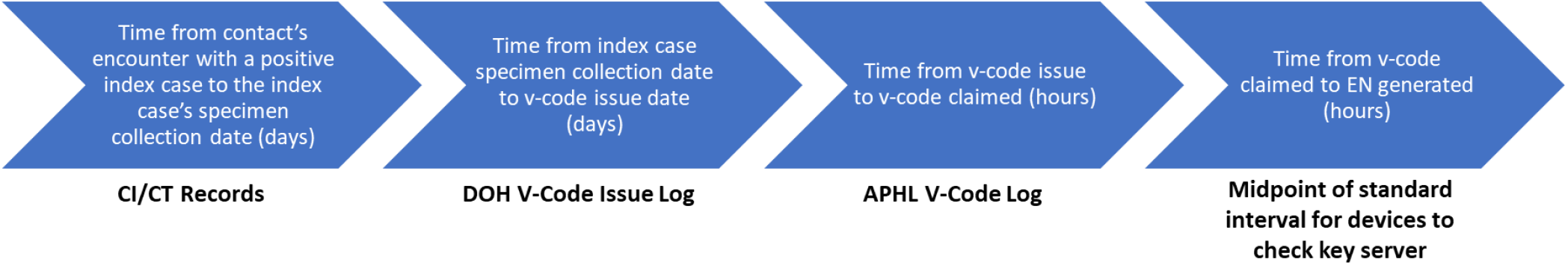
Estimating time from exposure encounter to receipt of EN.

#### Adherence to quarantine (iv)

To obtain an estimate of quarantine adherence, responses from the Protective Behavior Surveys were averaged, resulting in an estimated 53% quarantine adherence. This estimate takes into account that intention to and actual quarantine may differ and that survey respondents may be more willing to adhere to quarantine guidelines. A similar approach (averaging two survey estimates) was used to calculate a quarantine adherence rate of 45.5% by Wymant et al^12^.

#### Size of transmission chain from a single case (v)

Size of the transmission chain is a function of the number of cases reported during the study period. If *C*(*t*) is the number of cases, *C*, in WA at time *t*, the number of cases averted at time *T* because of a single transmission at time *t* is equal to 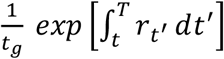, where *t*_*g*_ is the mean generation time (using the estimate *t*_*g*_ = 5.5, following methods used in Ferreti 2020)^21,22^ and *r*_*t*_ is the epidemic growth rate at time *t*. At a population level, if we assume: i) transmission of the virus outside of the state can be ignored; ii) the number of new cases will be small enough to not affect the exponential epidemic growth; iii) NPIs would be the same even without WA Notify; and iv) the growth rate will not change with a small number of additional cases, we obtain 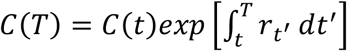 for any *t* < *T*. Therefore, instead of estimating the growth rate *r*_*t*_ which is related to the reproduction number and mean generation time, we can use the factor *C*(*T*)/*C*(*t*)*t*_*g*_ for each transmission at time *t*. The number of cases *C*(*t*) can be estimated from a 7-day moving average of new confirmed cases in WA State.

## Results

The modeling to estimate number of cases averted used variations of the parameter calculations presented above. The chosen parameters used reflect team and DOH congruence regarding most realistic variation in SAR and quarantine behavior within the bounds of the modeling approach. Fig. 2 illustrates the estimated number of COVID-19 secondary transmissions/cases averted at varying levels of SAR and quarantine effectiveness.

**Figure 2.**
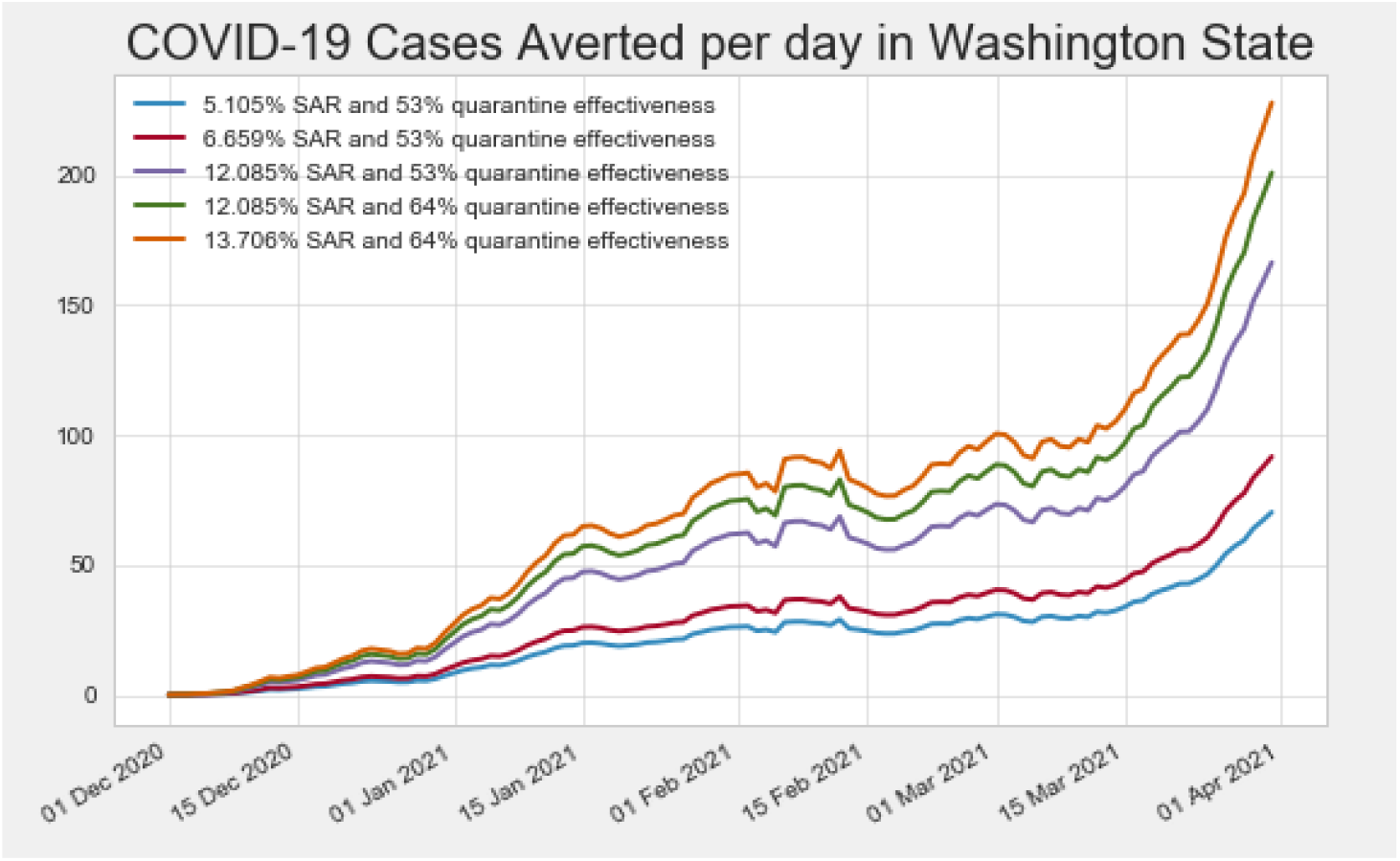
COVID-19 cases averted per day in WA State.

As illustrated, modeling estimates that between 11/30/2020 and 03/31/2021:

- Assuming 5.1% SAR and 53% quarantine effectiveness, 2636 cases are averted.
- Assuming 6.659% SAR and 53% quarantine effectiveness, 3439 cases are averted.
- Assuming 12.085% SAR and 53% quarantine effectiveness, 6240 cases are averted.
- Assuming 12.085% SAR and 64% quarantine effectiveness, 7536 cases are averted.
- Assuming 13.706% SAR and 64% quarantine effectiveness, 8547 cases are averted.

After conducting a sensitivity analysis (Figure 3), the estimated number of COVID-19 cases averted is 5,500 (central 95% range of sensitivity analysises 2,800-8,200). Additional details are included in Section I of the *Appendix*.

**Figure 3.**
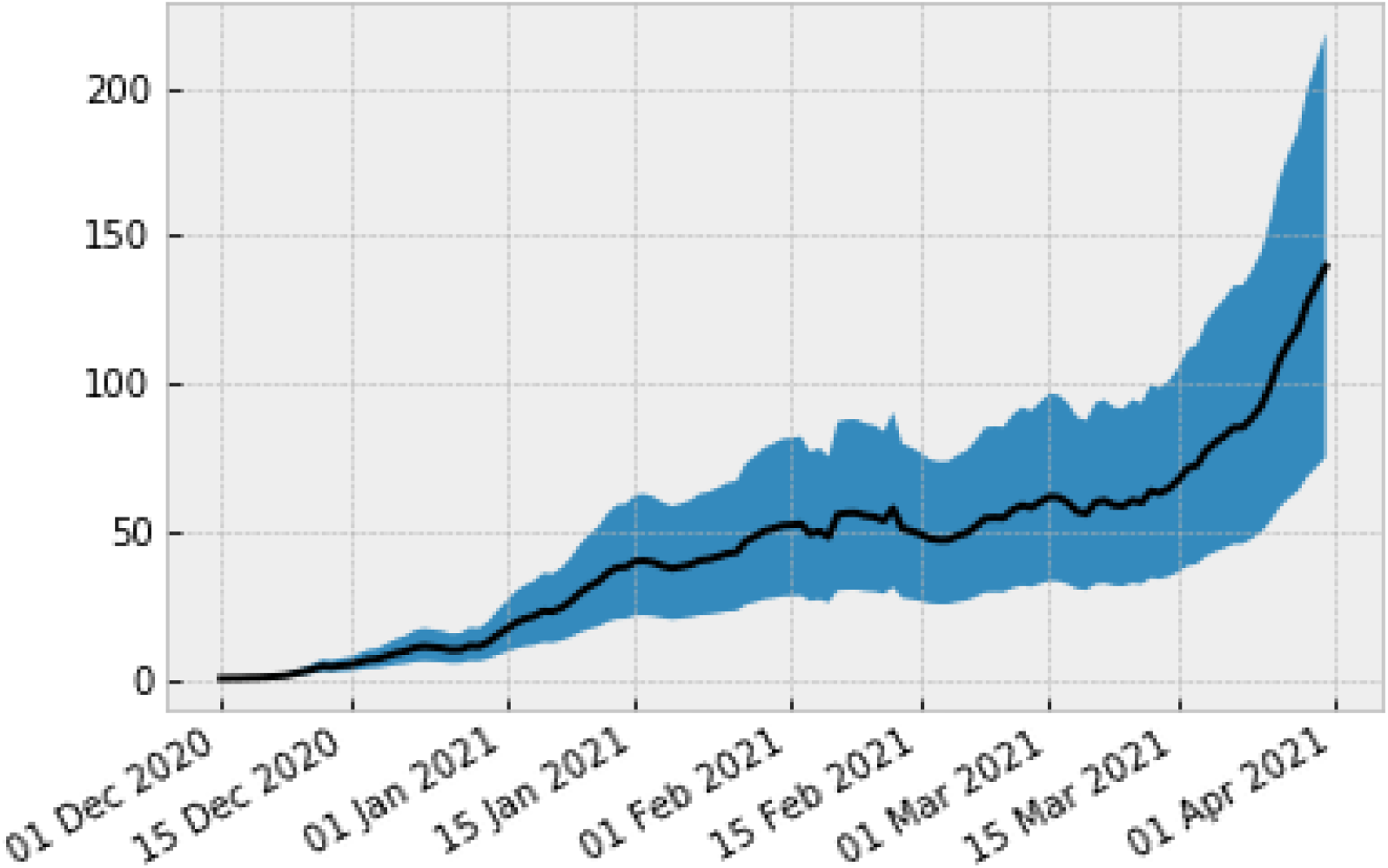
Sensitivity Analysis on COVID-19 Cases Averted Modeling.

Applying an estimated case fatality of 1.4% in WA State to the cases averted estimates,^16^ WA Notify saved 40-115 lives between November 30, 2020 and March 31, 2021.

## Limitations

There are several limitations to our work. First, privacy preservation of EN tools prevents directly observing ENs received and opened. The CI/CT dataset was extracted from DOH surveillance systems not intended for research, therefore we cannot explain all data missingness due to human error or variation in documentation protocols across PHAs. In addition, the CI/CT dataset only includes cases interviewed by LHJs that use the state’s primary CI/CT surveillance system and does not include CI/CT data from the LHJs that use their own systems. Lastly, as an observational study the counterfactual of the number of cases reported in WA without WA Notify available as an intervention cannot be compared to our modeling estimates.

## Discussion

This analysis is among the first to quantify the public health value of digital proximity-based EN tools for COVID-19 control in the U.S. Our results support the use of EN tools such as WA Notify as an evidence-based, non-pharmaceutical intervention for pandemic response. During our evaluation period, 10,084 WA Notify users anonymously reported their positive COVID-19 diagnosis, which generated an estimated 34,501 ENs, representing engagement among users in public health efforts to protect their communities. Most notably, our analysis demonstrates that adoption of WA Notify in Washington state contributed to COVID-19 mitigation efforts with an estimated 5,500 COVID-19 cases and approximately 77 deaths averted in Washington between November 30, 2020-March 31, 2021. Digital EN tools do not replace traditional CI/CT processes; both are vital to COVID-19 pandemic control efforts. However, our analysis suggests that WA Notify supplements traditional CI/CT outreach efforts in Washington and may reach a unique population of individuals unknown to the index case with 3.4 ENs generated per code compared to 2.7 contacts reported per confirmed index case through CI/CT.

As an anonymous, privacy-preserving tool there are limitations to understanding the userbase and how the tool may motivate users to engage in protective behaviors^23^. Our use of aggregate metrics from available public and de-identified data sources presents one strategy for conducting future evaluations that could examine additional impacts of WA Notify^24^. Further investigation into the user experience through surveys as well as the reach and timeliness of WA Notify compared to traditional CI/CT is warranted to improve our understanding of its impact on collective public health efforts.

GAEN represents a private-public partnership in technology to scale the communication of risk notification and supplement the tools PHAs have to support their communities for the control of infectious diseases. The partnership with private sector technology providers (Google, Apple and Microsoft) has resulted in a remarkable rapid transformation of a methodology from concept into large scale practice^25^. This includes the collaboration on both the creation of the API and the transition from custom application towards a more rapidly adopted shared technology EN Express^26^.

EN systems are just one tool among many (vaccines, CI/CT, masks, hygiene, social distancing, occupancy policies) that should be leveraged to contain the spread of COVID-19, and with the expanding evidence-base for their effectiveness, integrating tools like WA Notify into public health practice alongside other established interventions will be helpful for epidemic control in the future. As new variants emerge and non-essential travel bans are lifted, there is uncertainty about the future of the COVID-19 pandemic^27^ and a need to maintain effective pandemic control strategies. With digital EN tools like WA Notify being implemented worldwide, they could potentially be leveraged to strengthen measures against COVID-19 spread among travelers.

We view our findings as evidence of a new emerging paradigm for public health in which innovative technologies like WA Notify are utilized synergistically with other established NPIs like traditional CI/CT to produce multiplicative benefits for pandemic control. As we emerge from the COVID-19 pandemic, the application of digital proximity detection and anonymous ENs may shift to other public health emergencies in the future and strengthen community-clinical engagement with added functionality (e.g., requesting a test or reporting vaccination status)^28^.

## Data Availability

The data is not publicly available.

## Disclosures

### Authors’ contributions

CS, ZZ, BK, DR, GZ and JB contributed to study design, data collection and data interpretation; CS and ZZ conducted data analysis; All authors contributed to manuscript preparation and revision.

## Acknowledgements

We wish to thank our collaborators at the Washington State Department of Health, notably Amanda Higgins and Andrea King. We are grateful for the contributions of our team members in scoping data analysis plans, Daniel Lorigan, and conducting data analysis, Tiffany Chen, and greatly appreciate the time and feedback provided by Christophe Fraser and Luca Ferretti of the Oxford Big Data Institute on our approach and preliminary results.

