## Supplementary Appendix for "Early Epidemiological Evidence of Public Health Value of WA Notify, a Smartphone-based Exposure Notification Tool: Modeling COVID-19 Cases Averted in Washington State"

Affiliations:
[1]Department of Health Services, University of Washington
[2]Department of Statistics, University of Washington
[3]Washington State Department of Health
[4]Department of Epidemiology, University of Washington

### Supplementary Appendix

This appendix has been provided by the authors to give readers additional information about their work.

Table of Contents

1. WA Notify and the CI/CT Process in WA State
2. The WA State COVID-19 Landscape during the Modeling Time Period
3. Data Sources Used for Modeling Parameters
4. Supplementary Analyses Used for Parameter Estimation

1. WA Notify and the CI/CT Process in WA State

Figure S-1 describes the CI/CT interview workflow of three “types” of positive index case: a WA Notify user reporting a positive diagnosis (i.e., claims a code); a WA Notify user who receives an exposure notification (EN), gets tested, tests positive and is contacted for a CI/CT interview; and an individual who tests positive for COVID-19 but did not receive an EN.

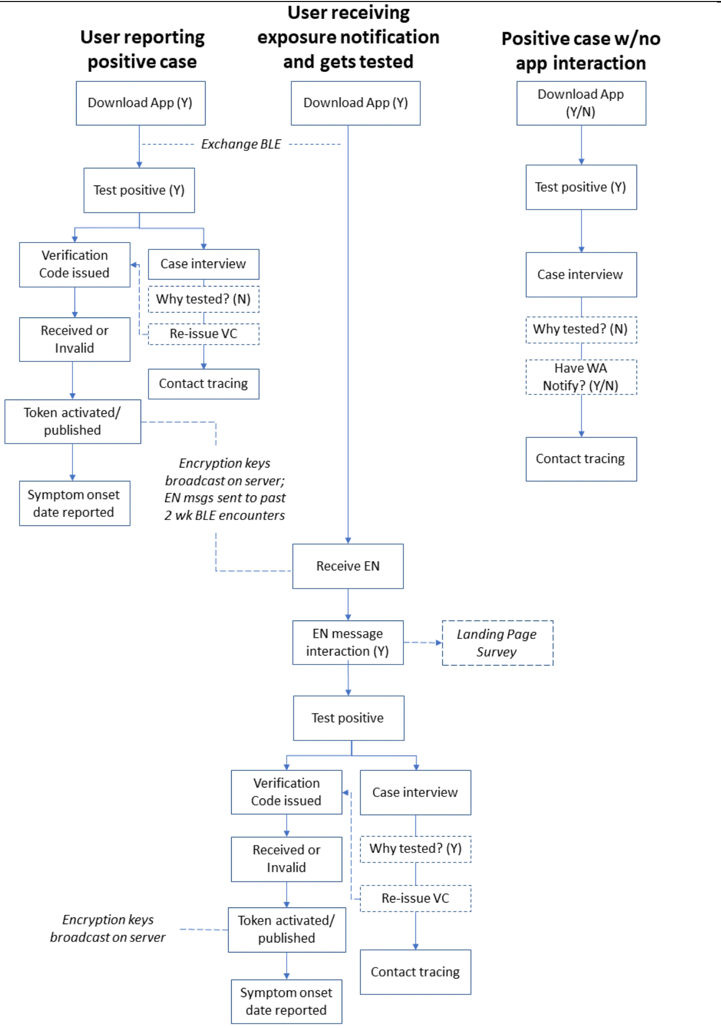

Figure S-1: Workflow Diagram: WA Notify and the CI/CT Interview Workflow in WA State

2. The WA State COVID-19 Landscape during the Modeling Time Period

Figure S-2 displays the weekly count of COVID-19 diagnoses reported in WA State during the study period. Figure S-3 displays the weekly count of hospitalizations and deaths reported in WA State during the study period.

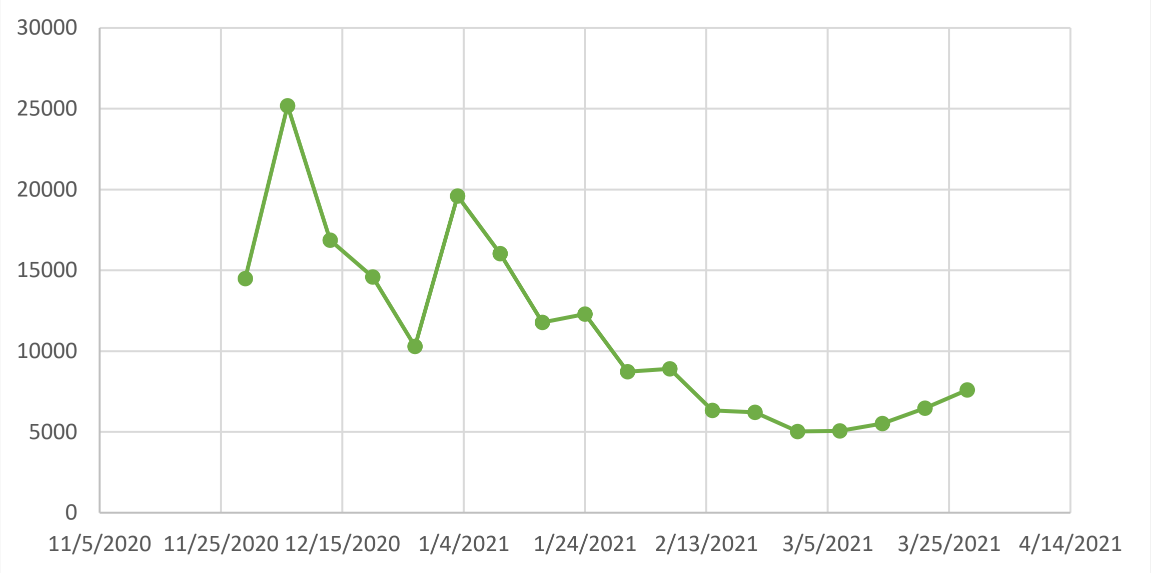

Figure S-2: Confirmed and Probable COVID-19 diagnoses (11/30/20-3/31/21)

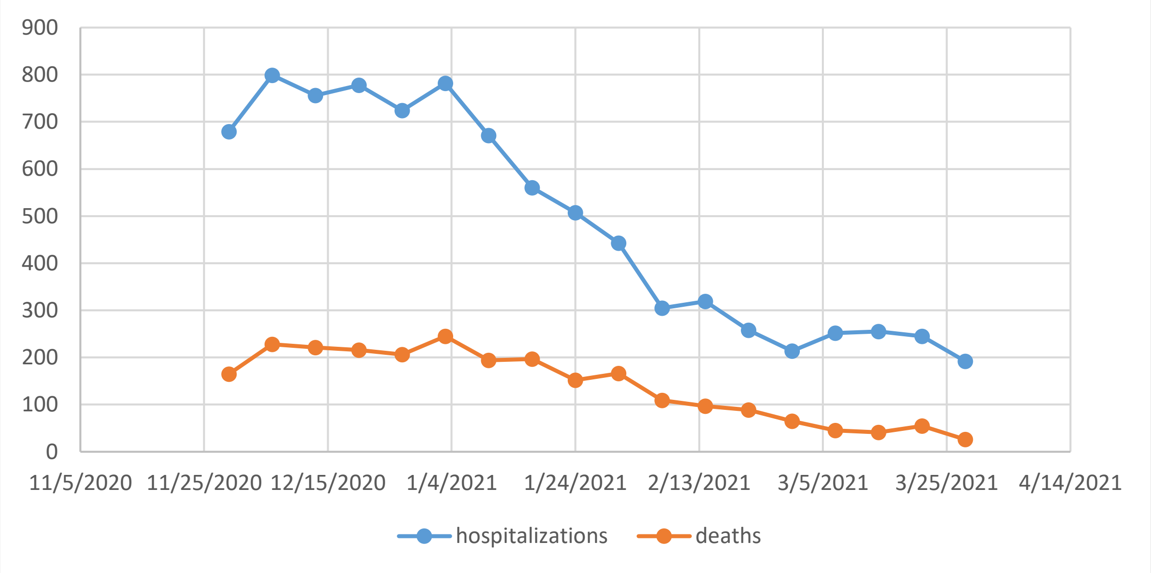

Figure S-3: COVID-19 Hospitalizations and Deaths (11/30/20-3/31/21)

3. Data Sources Used for Modeling Parameters

Table S-1 provides information about the metrics, timeline, and application of each of the following datasets used in the evaluation. Detailed descriptions of each dataset follow.

**Table S-1. Data Sources, Description and Utilization in Model**

| *Source* | *Timeframe* | *Data Pulled* | *Utilization in Model* |
|---|---|---|---|
| **DOH CI/CT Data** | 11/01/2020—03/19/2021 | Positive contacts among index cases | Secondary attack rate |
| | | Time from contact's exposure date to index case's specimen collection date | Delay of EN |
| | | Date of first successful contact tracing call | Delay of EN |
| | | Date of specimen collection | Delay of EN |
| **DOH Bulk Code Issue Log Data** | 01/12/2021—02/05/2021 | Time from specimen collection to code issue | Delay of EN |
| **APHL EN Verification Code Server Metrics** | 02/03/2021-03/31/2021 | Code claim "age" (in hours) distribution | Delay of EN |
| **ENPA Dashboard Data** | 02/09/2021-03/31/2021 | Number of notifications generated | Estimated number of ENs generated |
| | | Number of notifications opened | Estimated number of ENs generated |
| **DOH Landing Page "What to do next" Data** | 11/30/2020-03/31/2021 | Page hits each day | Estimated number of ENs generated |
| **Protective Behavior Surveys** | 01/30/2021-04/21/2021 | Proportion of respondents that intend to quarantine (Survey 1) and report quarantining (Survey 2) after receiving an EN | Adherence to quarantine |
| **WA State DOH Dashboard** | 11/30/2020-03/31/2021 | Cumulative and weekly cases | Fraction of transmissions prevented |

**DOH CI/CT Data**: The CI/CT process begins with a positive COVID-19 test result. When the lab test result is confirmed or probable, the index case, along with corresponding identifiers including demographics, addresses, and phone numbers, is reported to the Washington Disease Reporting System (WDRS) and then uploaded to the Case Risk Exposure and Surveillance Tool (CREST) system for CI/CT management. CREST was a new tool deployed in late November 2020 by DOH specifically for the capture of COVID-19 CI/CT data. The traditional follow-up as seen in Figure S-1, entails CI/CT teams attempting to reach all positive index cases. The DOH CI/CT Dataset includes de-identified records representing DOH Centralized Investigations and local health jurisdiction (LHJ) CREST users collected between 11/01/2020—03/19/2021. The dataset includes a total of 286,856 records, of which 213,634 are case investigation records and 73,222 are contact survey records. Records with a number included in the WDRS ID variable were defined as positive cases. There were 54,990 case investigation records and 50,074 contact survey records that were completed post-launch of WA Notify (11/30/20). There were 49,488 contact survey records that had a non-missing index “Source ID”, which means that the identifier for the index case was recorded, allowing us to link the contact survey record with the index case. Matching the CREST “Source ID” between case investigation records with contact survey records, this sample includes 18,616 case investigation records who reported at least one contact, representing an average of 2.7 contacts per index case.

**DOH Bulk Code Issue Log Data:** Every day, epidemiologists at DOH pull WDRS lab records (including phone number and specimen collection date) with a “create date” of the prior day into the Bulk Issue Log . Phone numbers are uploaded for batch (SMS) issuance of verification codes confirming a positive COVID-19 test result to WA Notify users. Confirmation of SMS text delivery is recorded in a log. This information was used to estimate the delay in time from lab reporting to code issuance.

**APHL Verification Code Server Metrics:** In order for WA Notify users to report a positive diagnosis in WA Notify, they must receive a verification code through the APHL server. Aggregate, de-identified data of verification code deployment and claimed codes are made available to DOH. One metric is the distribution of time between issuance of a new code to the user claiming the code.

**ENPA Dashboard Data**. The Exposure Notifications Private Analytics (ENPA) Dashboard developed by The MITRE Corporation allows PHAs to access aggregated, anonymous data from users who opted-in to report analytics when they activated WA Notify. Data regarding ENs received, opened, or dismissed were available for the evaluation beginning 02/09/2021, representing a total of approximately 28,958,065 device-days contributing to the aggregated analysis. Using the percentage of users who opted-in to share analytics (1.03%-3.75%) multiplied by the total number of iOS and Android devices with WA Notify installed at the time (1,821,230-1,881,447), indicated there were 10,741 notifications generated and 5,215 notifications opened between 03/01-03/31/2021.

**DOH Landing Page "What to do next" Data**. The EN message delivered to a WA Notify user’s phone provides a link to a hidden Landing Page on the WA State DOH website that provides guidance (testing, protective behaviors, quarantine) after learning of a potential exposure to a person who tested positive for COVID-19. The link to this page is only accessible to WA Notify users who tap the EN message link. Between 11/30/2020—03/31/2021, there were 16,748 Landing Page hits.

**Protective Behavior Surveys**. In late January 2021, an online, anonymous survey (Survey 1) was added to the Landing Page. This brief survey included questions regarding intent to seek testing and to engage in protective measures (quarantining/staying home for 10 or 14 days, avoiding public places for 10 or 14 days, and/or staying away from others in their household). Respondents selecting any of these protective measures were classified as “intending to quarantine”. Between 01/26 and 04/21/2021, of the 1,132 responses to this question, 475 (42%) reported intending to quarantine. Survey 2 was distributed to respondents who were willing to receive a follow-up survey in approximately two weeks. This survey asked if the respondent had engaged protective measures. Of the 219 follow-up respondents, 140 (64%) reported having engaged in some form of quarantine behavior after receiving an EN between 01/26 and 04/21/2021.

4. Supplementary Analyses Used for Parameter Estimation

A. Verification Codes Issued and Claimed.
The APHL server provides data regarding the variety of ways codes can be issued (API based, manual via CI/CT interview, or automated SMS), the number of codes claimed and the age distribution in hours of codes claimed since the time the code was issued (available starting 2/3/21). The flexibility in code deployment introduces ambiguity in code issuance data for the evaluation. In addition, the same individual may also receive a code more than once, therefore the precise number of codes claimed is a more reliable metric compared to codes issued. Between 11/30/2020—01/10/2021, 1,952 codes were claimed, representing 1.8% of all positive cases reported in WA State. As seen in Fig. S-4, following the implementation of a bulk SMS code deployment on 01/11/2021 and 03/31/2021, 8,132 codes were claimed, increasing the reach of codes claimed to 9.6% of all positive cases when bulk issue was live. Between 11/30/20-3/31/21 there were a total of 101,990 codes issued and 10,084 were claimed (9.9%), representing 5.1% of all positive cases reported during the study period.

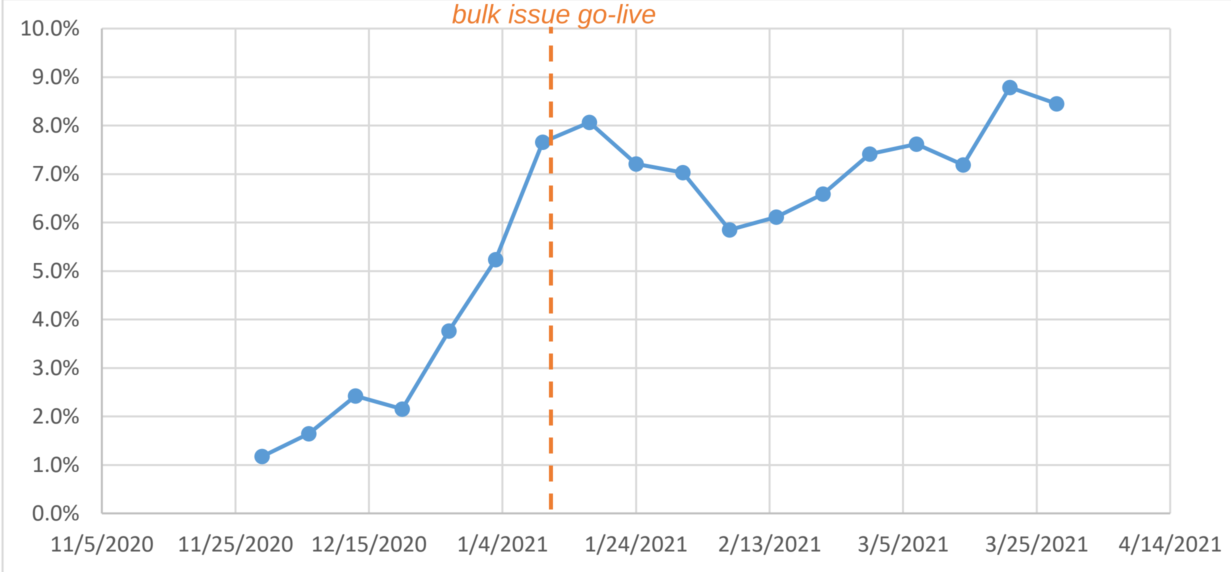

Figure S-4: Proportion of verification codes claimed among all reported COVID-19 diagnoses (11/30/20-3/31/21)

B. Assessment of Fit: Estimate of Number of ENs Generated.
A time series analysis (Fig. S-5) to assess the fit of the estimated signal based on the number of page hits compared to the number of notifications reported in ENPA during March 2021 (both smoothed to a 7-day moving average) yields a Pearson R = 0.98 over the time period 3/1-3/31, suggesting a good fit.

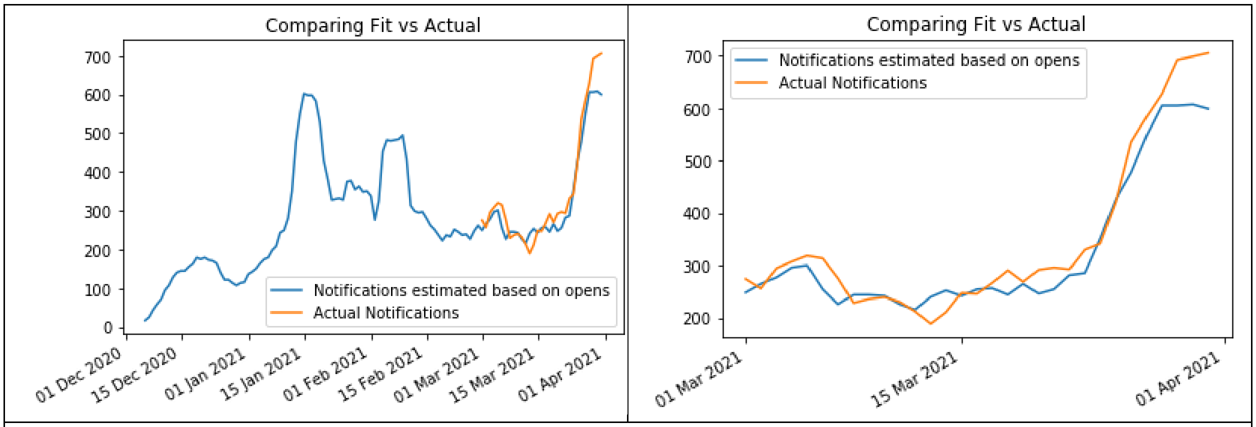

Figure S-5. Time-series analyses to assess fit.

C. Standard Deviation Propagation for Secondary Attack Rate
Let $SA_i$ be the count (expected value) on day $i$ of opted-in devices that verified a code and had received an exposure notification in the past 14 days. And $std_{SA_i}$ be the standard deviation of that count due to Gaussian noise injected by differential privacy. Let $Notify_i$ be the count (expected value) of opted-in devices that received a notification on day $i$. And $std_{notify_i}$ the standard deviation of that count due to Gaussian noise injected by differential privacy.
Let $\mathcal{D}$ be the set of dates that we are calculating these numbers. The expected value of SAR using data from $\mathcal{D}$ is given by:

$$\mathrm{SAR} = \frac{\sum_{i \in \mathcal{D}} SA_i}{\sum_{i \in \mathcal{D}} Notify_i}$$

Assume noises are independent Gaussian, the standard deviation of $SA = \sum_{i \in \mathcal{D}} SA_i$ is $\mathrm{std}_{SA} = \sqrt{\sum_{i \in \mathcal{D}} std_{SA_i}^2}$ and similarly. the standard deviation of $\mathrm{Notify} = \sum_{i \in \mathcal{D}} Notify_i$ is $\mathrm{std}_{Notify} = \sqrt{\sum_{i \in \mathcal{D}} std_{notify_i}^2}$.
By uncertainty propagation,

$$std_{SAR} = \frac{SA}{Notify}\sqrt{\left(\frac{std_{SA}}{SA}\right)^2 + \left(\frac{std_{Notify}}{Notify}\right)^2}$$

With a 68% confidence interval, the interval $SAR \pm std_{SAR}$ will cover the true secondary attack rate.

As of Washington's ENPA data, for $\mathcal{D}$ from 02/09/2021 to 05/04/2021, the interval we get for SAR is:

$$4.65\% \pm 0.973\%$$

For $\mathcal{D}$ from 03/01/2021 to 05/04/2021, the interval we get for SAR is:

$$5.12\% \pm 0.979\%$$

D. Time from Date of Contact's Exposure to the Index Case's Specimen Collection Date.

Included in the DOH CI/CT Dataset are variables utilized to estimate the delay from exposure to EN. Within the dataset there were 17,426 records with a value included for "Specimen collection date - Date of Exposure contact" of which there were 17,268 observations between -14 and 14 days, with a mean delay from the exposure encounter to specimen collection of 2.064 days, as illustrated in Fig. S-6.

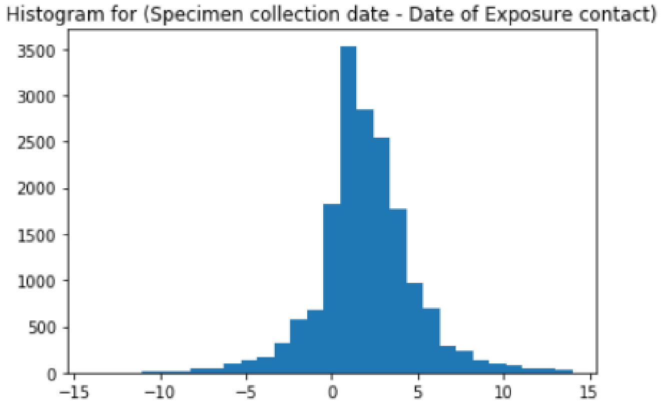

Figure S-6. Time from date of exposure to specimen collection data.

E. Time from Index Case's Specimen Collection Date to Code Issued.

Looking at the time between date of specimen collection to date of code issuance in the logs of bulk SMS deployment, 24,305 records out of 25,205 indicated a code was issued within 14 days (3.6% of the records indicated code issuance over 14 days), with a mean of delay of 3.4 days, as seen in Fig. S-7.

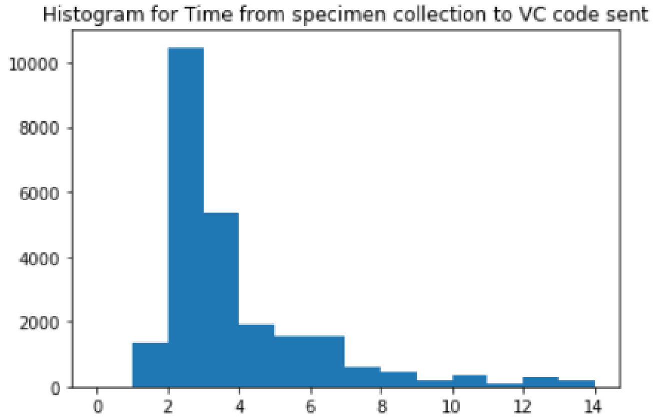

Figure S-7. Time from date of specimen to code issued.

F. Time from Code Issued to Code Claimed.

The APHL server tracks the time from code issue to code claimed in hours. This metric was available starting 2/2/21. The majority (72.2%) of codes claimed were claimed within 1 hour of issue (Fig. S-8).

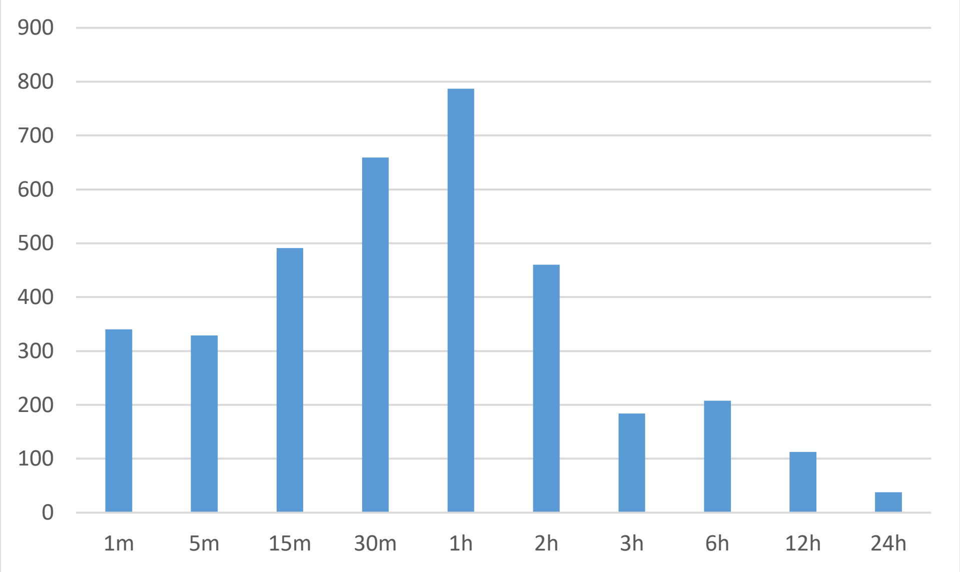

Figure S-8. Count of codes claimed by age of code from issue.

G. Time from Code Claimed to EN Received.
This estimate is based on the midpoint of the standard interval for GAEN-based systems to have devices check the key-server, which is 4 hours.

H. Evaluation of the Transmissions Prevented Distribution.
We use the Monte Carlo method to evaluate the unknown distribution of the observations and estimate the proportion of transmissions that can be prevented. A Monte Carlo sampling was applied to two segments of observations a) time from the date of a contact's exposure to the date of the index case's specimen collection data provided in the CI/CT dataset, as well as b) time from specimen collection to code issue provided in the DOH bulk issue logs. First, a sample time $x$ is taken from a) "specimen collection date to date of Exposure contact" from the CI/CT data, then a sample time $y$ is taken from b) "Specimen collection date to code sent date" from the bulk issue logs. The number of days from "contact's exposure to index case's code claim to EN generated" can be calculated as $x+y+0.167$ (0.167 days from code verified to notification sent). The corresponding percentile of the Cumulative Distribution Function (CDF) of the Weibull distribution was used to calculate the average time delay over multiple samples.

Applying this simulation (Fig. S-9) indicates approximately 51.5—52.3% of transmissions can be prevented by receipt of ENs. Among the observations used in the Monte Carlo sampling, 3.6% and 0.8% of the data fall outside of the 14-day period for which an receipt of an EN would have utility. Omitting these data, we conclude that (1-3.6%-0.8%)*51.9%=49.62% of transmissions could be prevented. The distribution of transmission time is modelled by a Weibull distribution (shape=3.2862, scale=6.1244)[26]. The cumulative density of transmissions prevented at 1.5 days is approximately 0.01, 0.40 at 5 and 0.95 at 8.5—in other words, WA Notify was preventing 99% of transmissions if the delay from exposure to EN was less than 1.5 days, 60% of transmissions if the delay from exposure to EN was 5 days, and only 5% of transmissions if the delay was 8.5 days.

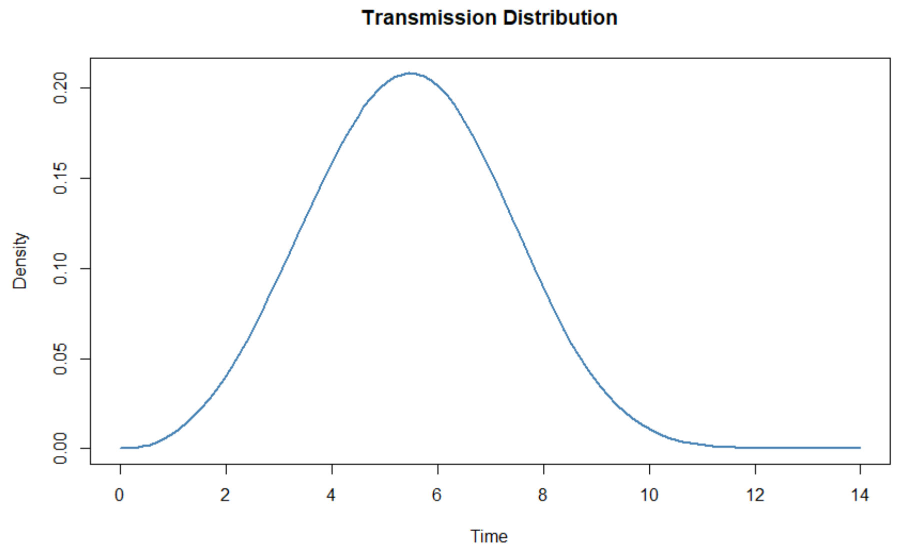

Figure S-9. Estimated transmission distribution over 14 day period

I. Sensitivity Analysis of the Modeling Results.
Following the approach used in Wymant et al 2021, we consider four sources of uncertainty for the sensitivity analysis on the modelling results:

1. Uncertainty about the effectiveness of quarantine: we assume a uniform distribution between 53%-64%.
2. Uncertainty about the SAR: we assume a uniform distribution between the values of 5.105% and 13.706% found by varying the delays involved in estimating SAR.
3. Uncertainty about the generation time distribution, which corresponds to the distribution of the timing of transmission from exposure and the distribution of the delay from exposure to notification. We assume that the parameters of the Weibull distribution are normally distributed (SD=0.75 for the shape, SD=0.5 for the scale), which represent a good approximation of the uncertainty from Ferretti et al., *Science* 2020.
4. Uncertainty about the delays from exposure to notification: we assume that the uncertainty is captured by the Monte Carlo Sampling described in Section H above. We combined the delay and generation time parameters by choosing shape and scale from our previous normal distributions and then compute the Monte Carlo to estimate the proportion of transmissions that can be prevented.

Given the large number of notifications, we assume that the relative uncertainty on the number of notifications is small. In addition, the uncertainty propagation on the ENPA data used to calculate the error on the lower bound SAR estimate shows that the standard deviation for the number of notifications is <1% of the total number of notifications, and that the same is true for the uncertainty about the expected size of a transmission chain, hence we neglect these uncertainties. After including these uncertainties in the modelling, we compute sensitivity intervals as the 2.5% and 97.5% quantiles for each estimate in the model.
